# Chloroquine, hydroxychloroquine, and COVID-19: systematic review and narrative synthesis of efficacy and safety: Systematic review of (hydroxy)chloroquine efficacy and safety

**DOI:** 10.1101/2020.05.28.20115741

**Authors:** Michael Takla, Kamalan Jeevaratnam

## Abstract

**Background:** The COVID-19 pandemic has required clinicians to urgently identify new treatment options or the repurposing of existing drugs. Several drugs are now being repurposed with the aim of identifying if these drugs provide some level of disease resolution. Of particular interest are chloroquine (CQ) and hydroxychloroquine (HCQ), first developed as an antimalarial therapy. There is increasing concern with regards to the efficacy and safety of these agents. The aims of this review are to systematically identify and collate studies describing the use of CQ and HCQ in human clinical trials and provide a detailed synthesis of evidence of its efficacy and safety.

**Methods and Findings:** Searches for (“COVID” AND “chloroquine”[title/abstract] AND “outcomes”[full text]) and two (“COVID” AND “hydroxychloroquine”[title/abstract] AND “outcomes”[full text]) yielded 272 unique articles. Unique articles were manually checked for inclusion and exclusion criteria and also subjected to a quality appraisal assessment. A total of 19 articles were included in the systematic review. Seventy-five percent of observational studies employing an endpoint specific to efficacy recorded no significant difference in the attainment of outcomes, between COVID-19 patients given a range of CQ and/or HCQ doses, and the control groups. All clinical trials and 82% of observational studies examining an indicator unique to drug safety discovered a higher probability of adverse events in those treated patients suspected of, and diagnosed with, COVID-19. Seventy-five percent of the total papers focusing on cardiac side-effects found a greater incidence among patients administered a wide range of CQ and/or HCQ doses, with QTc prolongation the most common finding, in addition to its consequences of VT and cardiac arrest. Of the total studies using mortality rate as an end-point, 60% reported no significant change in the risk of death, while 30% showed an elevation, and 10% a depression, in treated relative to control patients.

**Conclusion:** The strongest available evidence suggests that, relative to standard in-hospital management of symptoms, the use of CQ and HCQ to treat hospitalised COVID-19 patients has likely been unsafe. At the very least, the poor quality of data failing to find any significant changes in the risk of VT should preclude definitive judgment on drug safety until the completion of high-quality randomised clinical trials.

## 1.0 Introduction

In the final week of December 2019, the Hubei Integrated Chinese and Western Medicine hospital in Wuhan, reported a clustered point-source outbreak of pneumonia [1], of unknown viral origin. Within 30 days, the rapid geographic expansion of the disease, which the International Committee on Taxonomy of Viruses later coined Coronavirus Disease 2019 (COVID-19) [2], implied propagation by human-to-human transmission. On March 11 2020, the World Health Organisation (WHO) designated COVID-19 a pandemic [3]. As of May 26 2020, COVID-19 has been confirmed as the cause of 5,508,904 cases and 346, 326 deaths [4] globally.

In the absence of specific antiviral pharmacotherapy, the repositioning of existing drugs represents an attractive clinical option. Selecting which drugs to repurpose, however, hinges on the compatibility of their mechanisms of action with the disease progression of severe acute respiratory syndrome (SARS), and with the biology of the recently emerged pathogenic agent that causes it.

With a likely evolutionary origin in bats [5], the novel (beta)-coronavirus, SARSCoV-2, probably acquired the ability to zoonotically infect humans via natural selection of the receptor-binding domains of its spike (S) proteins in an intermediate mammalian host [6]. Indeed, compared to SARS-CoV-1, the highly homologous [7] coronavirus responsible for the SARS pandemic [8], the S protein has a 10–20-fold greater affinity for the ACE2 receptor [9] predominantly expressed by pulmonary and intestinal epithelia and vascular endothelia [10]. In fact, *in silico* analysis has demonstrated that the expression of sialic acid further facilitates viral entry, whereby binding of human gangliosides impairs inhibitory interactions between the S protein and the plasma membrane [11]. The resulting receptor-mediated endocytosis precedes endosomal cathepsin and TMPRSS2-mediated [12] cleavage of the S protein, permitting fusion of the viral lipid envelope and human vesicular membrane, whereupon RNA entry into the cytosol enables viral replication, maturation [13], and budding. The initial innate immune response to the subsequent dissemination of SARS-CoV-2 throughout the patient’s extracellular fluid elicits a wave of pro-inflammatory cytokines, including IL-1(beta), IL-6, and TNF-(alpha), that recedes upon lymphopenia, only to return at higher concentrations in a cytokine storm [14] that predisposes to a potentially lethal acute respiratory distress syndrome (ARDS). Additionally contributing to the mortality of critically ill COVID-19 patients is the significantly elevated incidence of often pulmonary thromboembolic events [15][16][17].

With antiviral [18][19], anti-inflammatory [20], and anti-thrombotic [21][22][23][24] effects, chloroquine (CQ), and its less oculotoxic [25][26] derivative, hydroxychloroquine, (HCQ) were among the first drugs selected for repurposing to treat COVID-19 patients. However, the ability of 4-aminoquinolones to prolong the QT interval [27][28] increases the risk of *de novo* ventricular tachyarrhythmias (VTs), calling into question their cardiac safety [29].

Here, we systematically review existing clinical trial data to provide a detailed synthesis of evidence for the efficacy and safety of CQ and HCQ. We also aim to clarify if the use of such drugs in COVID-19 patients in the absence of rigorous evidence may not only have had little efficacy, but, owing to their lack of (especially cardiac) safety, may have been responsible for excess mortality.

## 2.0 Review Methodology

### 2.1 Objectives

This systematic review seeks to clarify the strength of evidence for the relative efficacy and safety of CQ and HCQ treatment in patients suspected of, and diagnosed with, COVID-19.

### 2.2 Methods

In line with the PICOT format [30] of framing subjects for clinical research, this study centres on answering the question: ‘In patients suspected of, and diagnosed with, COVID-19, how efficacious and safe are CQ and HCQ, relative to standard symptomatic care?’

The subsequent elaboration of the systematic review and narrative synthesis adhered to the Preferred Reporting Items for Systematic Reviews and Meta-Analyses (PRISMA) guidelines [31] for evidence-based assessment of published research.

### 2.3 Search Strategy

In light of the current global health emergency, and the requisite rapid turnover of publications to meet the consequently urgent need to obtain and analyse the results that they present, several authors have resorted to the use of preprint servers to disseminate their findings. Despite the evident shortfalls inherent in referring to data, whose quality has not been peer-reviewed, the present paucity of published original research on the efficacy and safety of CQ and HCQ in COVID-19 patients demands exceptional measures. As such, this systematic review will take into account non-peer-reviewed work, provided that they have been submitted to a preprint server, where they are available in open-access form. Nevertheless, given its focus on data quality, this review will make unambiguous every instance in which data from such sources are used.

Therefore, on May 26 2020, MEDRXIV, along with PubMed, Web of Science and Embase, acted as the databases for the initial search of items relevant to the PICOT-formatted question. The preliminary use of the search terms “COVID”, “chloroquine”, and “hydroxychloroquine” yielded a large number of results that bore little relevance to the research topic. Combining such terms into phrases – *“COVID” AND “chloroquine”*, and *“COVID” AND “hydroxychloroquine”* in the title or abstract – and requiring the term *“outcomes”* anywhere in the full text considerably focused the responses. The subsequent application of identical phase stenography to each database ensured internal consistency.

### 2.4 Search Attrition Criteria

The aim of this review being to establish the weight of evidence for the use of a therapy in patients, data able to answer such a question must derive from primary research. Moreover, owing to the international extent of the present health crisis, any imposition of an original language requirement would exclude useful and otherwise rare resources. As such, following the collation of items in EndNote and the removal of duplicates, application of these criteria excluded unique items for which there was either no English version or no original data.

Screening of the resulting papers against the criteria established by the PICOT-formatted question – namely, the requirement that data be collected from COVID-19 patients treated with CQ and/or HCQ – included only controlled trials and observational studies.

### 2.5 Article Processing and Selection

Having applied the exclusion and inclusion criteria to all search results and removing duplicates at all stages where necessary, two investigators independently reviewed the final repertoire of studies.

### 2.6 Quality Appraisal

Rather than merely verifying the relevance and scope of the material in the final library, holistic analysis of each item of research in line with the framework set out in the Checklist of Review Criteria provided by the Task Force of Academic Medicine and GEARIME committee [32] ensured stringent appraisal of study quality.

Indeed, the identification of – among other facets of robust research – appropriate study design, statistical analysis, and quality control (Table 1) permitted only papers with sufficient scientific merit to pass onto the data extraction stage.

**Table 1:**
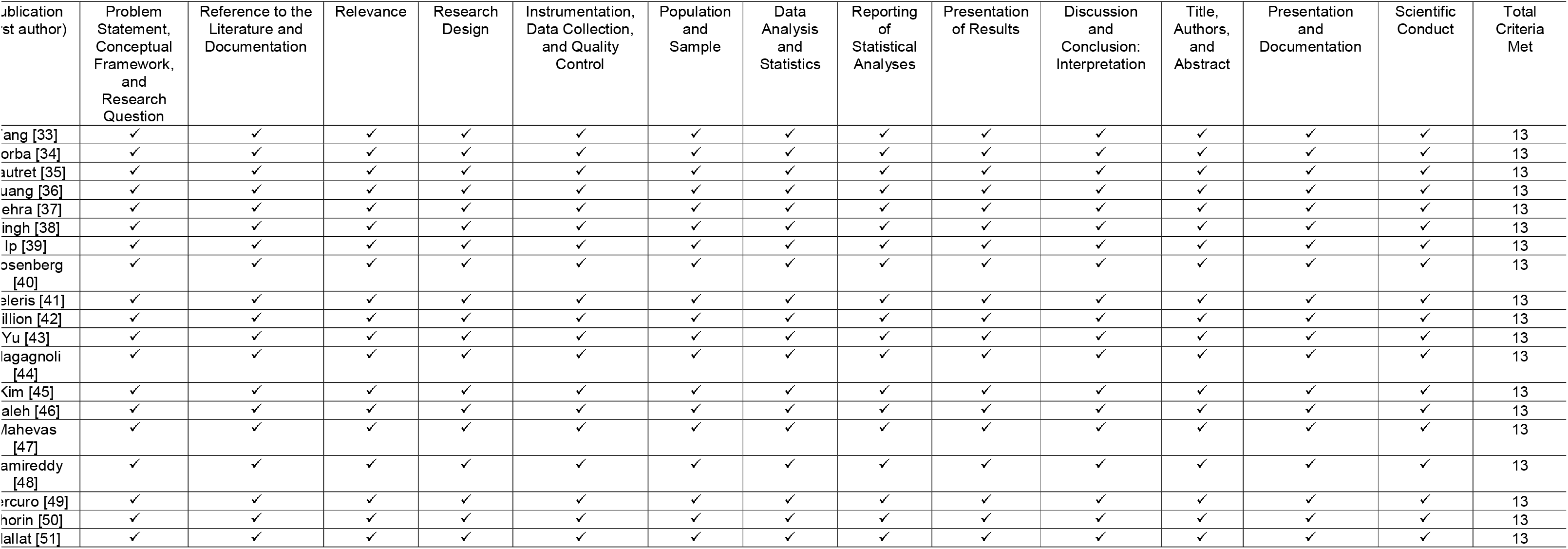
Quality appraisal of the 19 papers passing through search attrition.

### 2.7 Data Extraction

Among the research items constituting the final library for analysis, there exists a wide variation in study design, results, and, crucially, the extent to which each distinct aspect of the PICOT-formatted question is answered. As such, a specialised data extraction table collates and summarises the most important information in every paper (Table 2). In particular, emphasis on the different sample sizes and structures, doses of drug used, primary and/or secondary outcomes and overall design limitations, facilitates both clarity and caution when making comparisons between the sets of results presented.

**Table 2:**
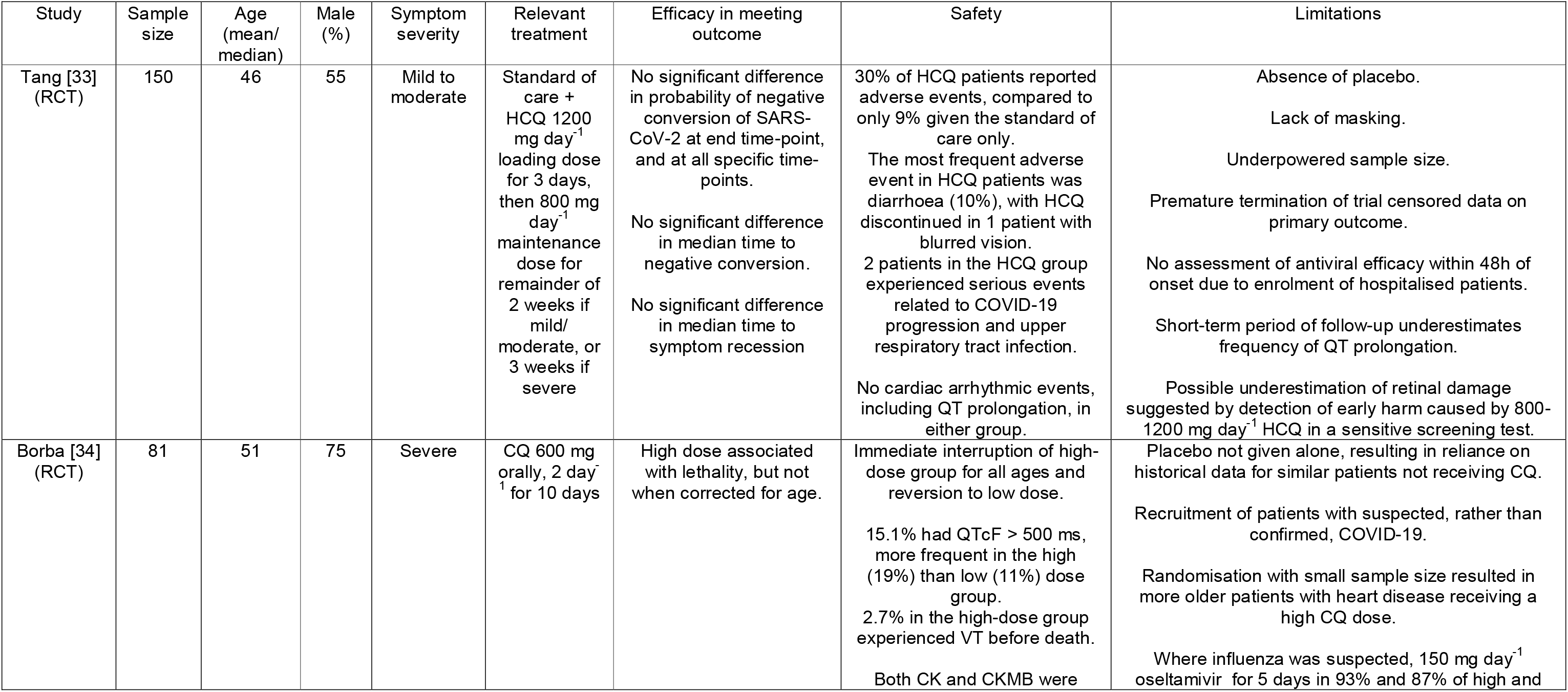
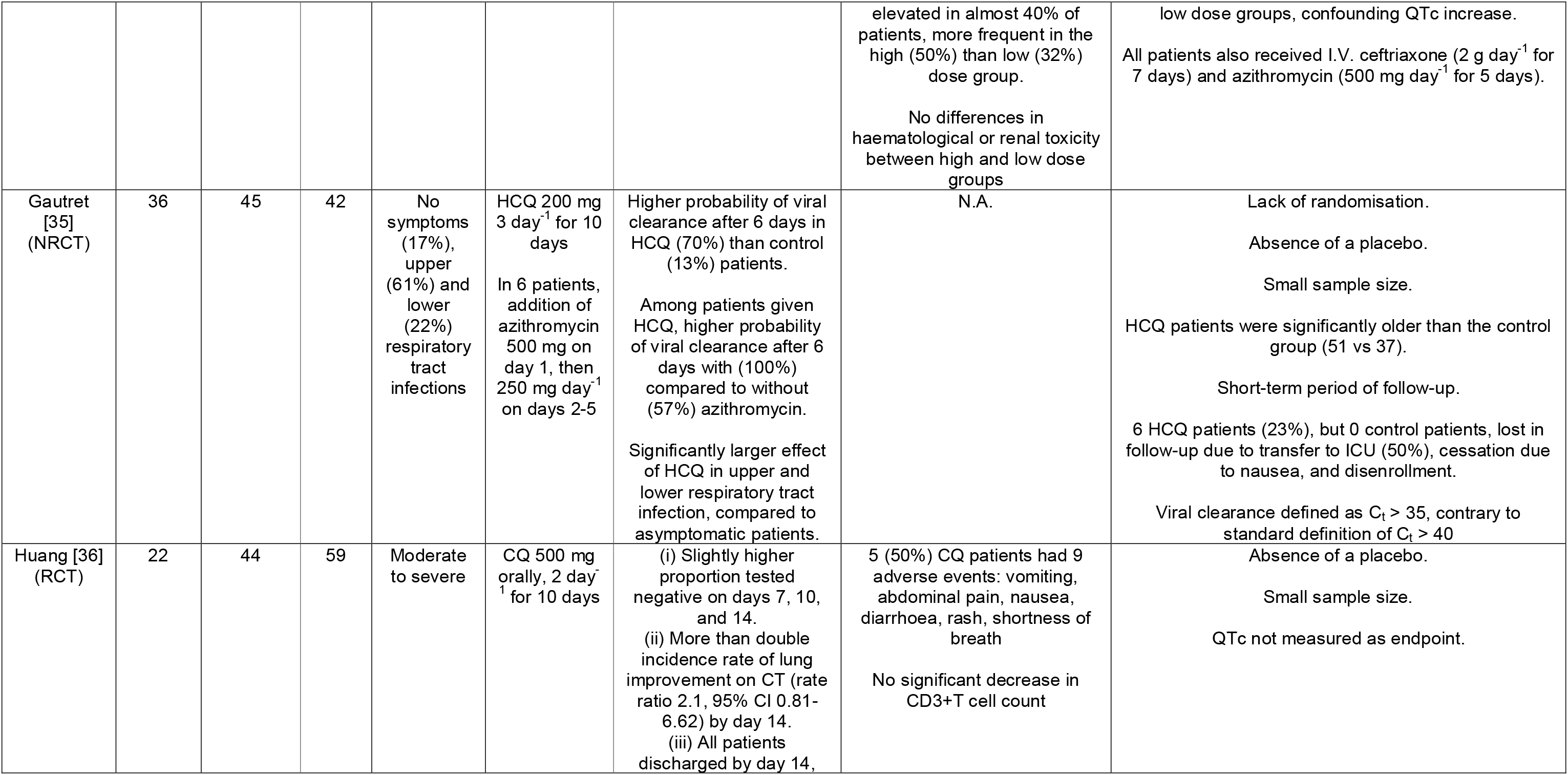
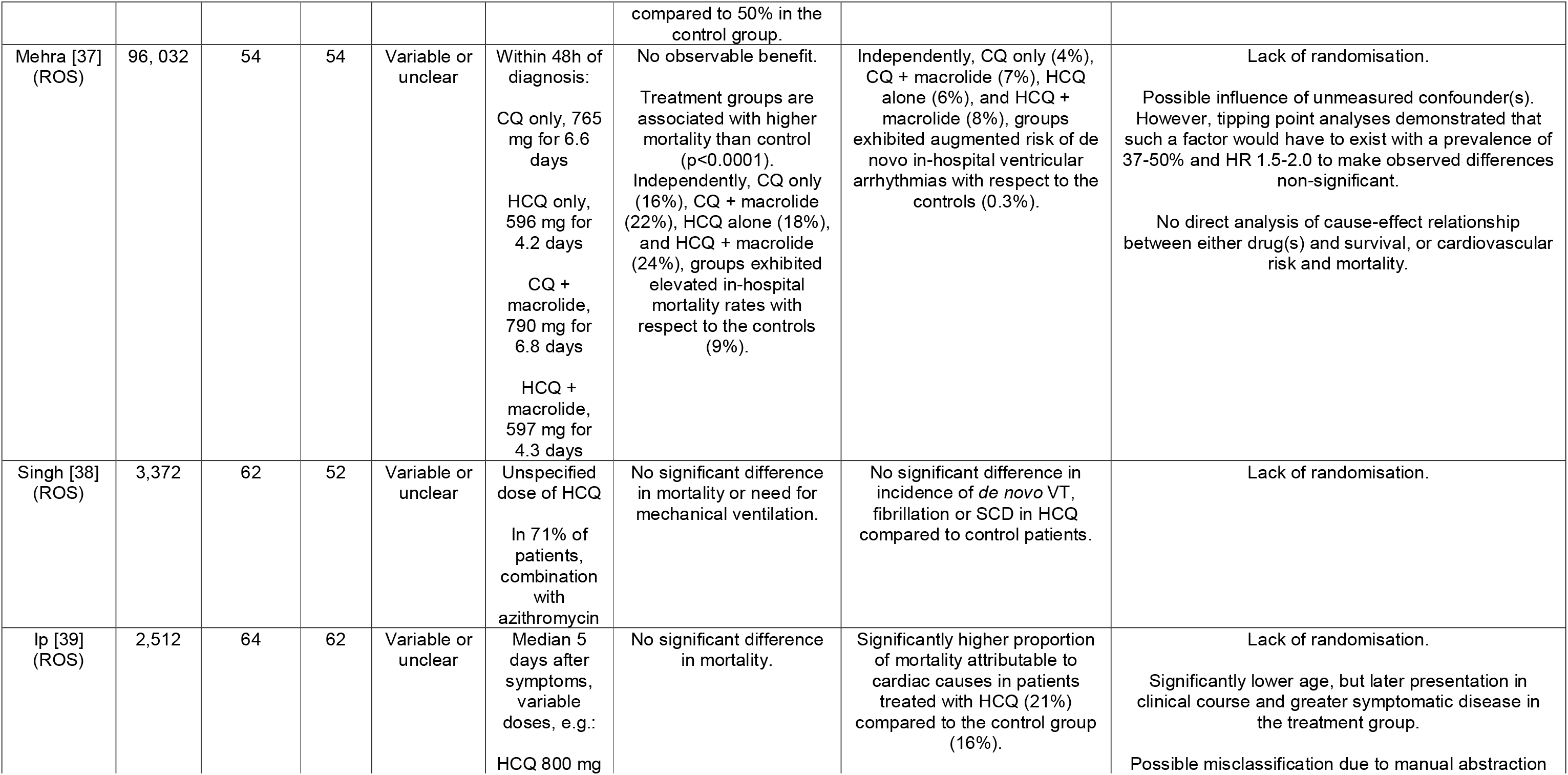
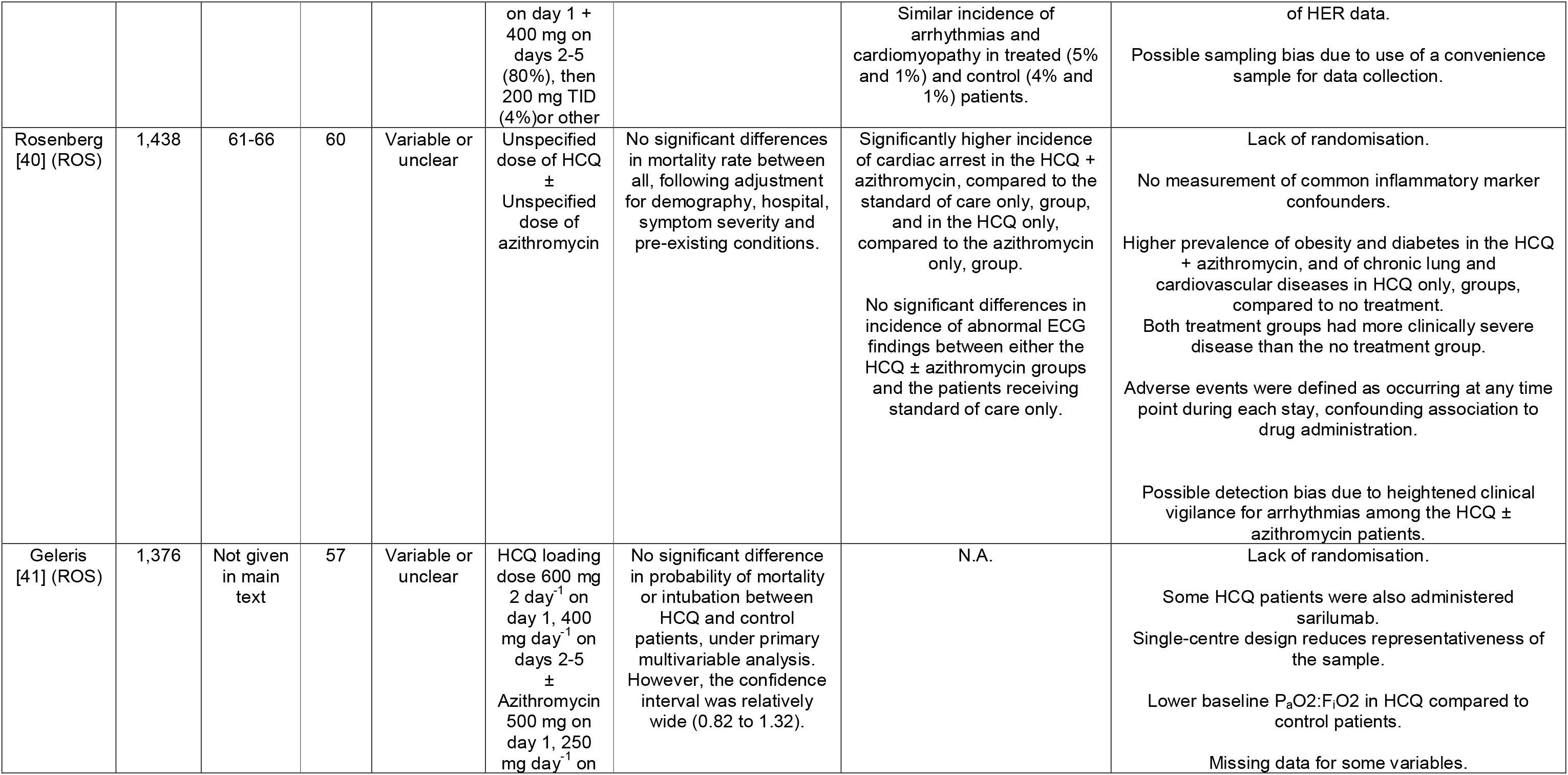
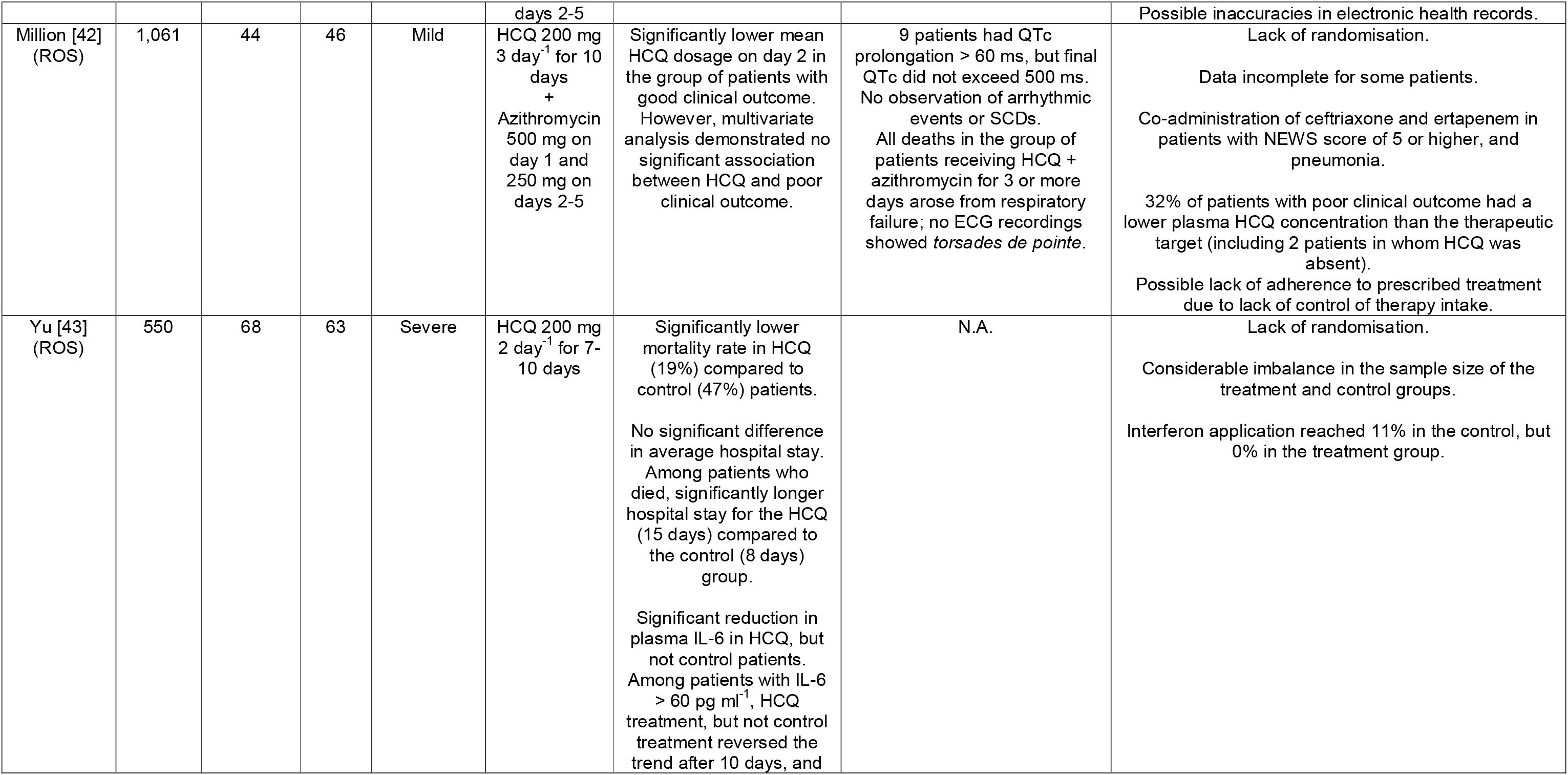
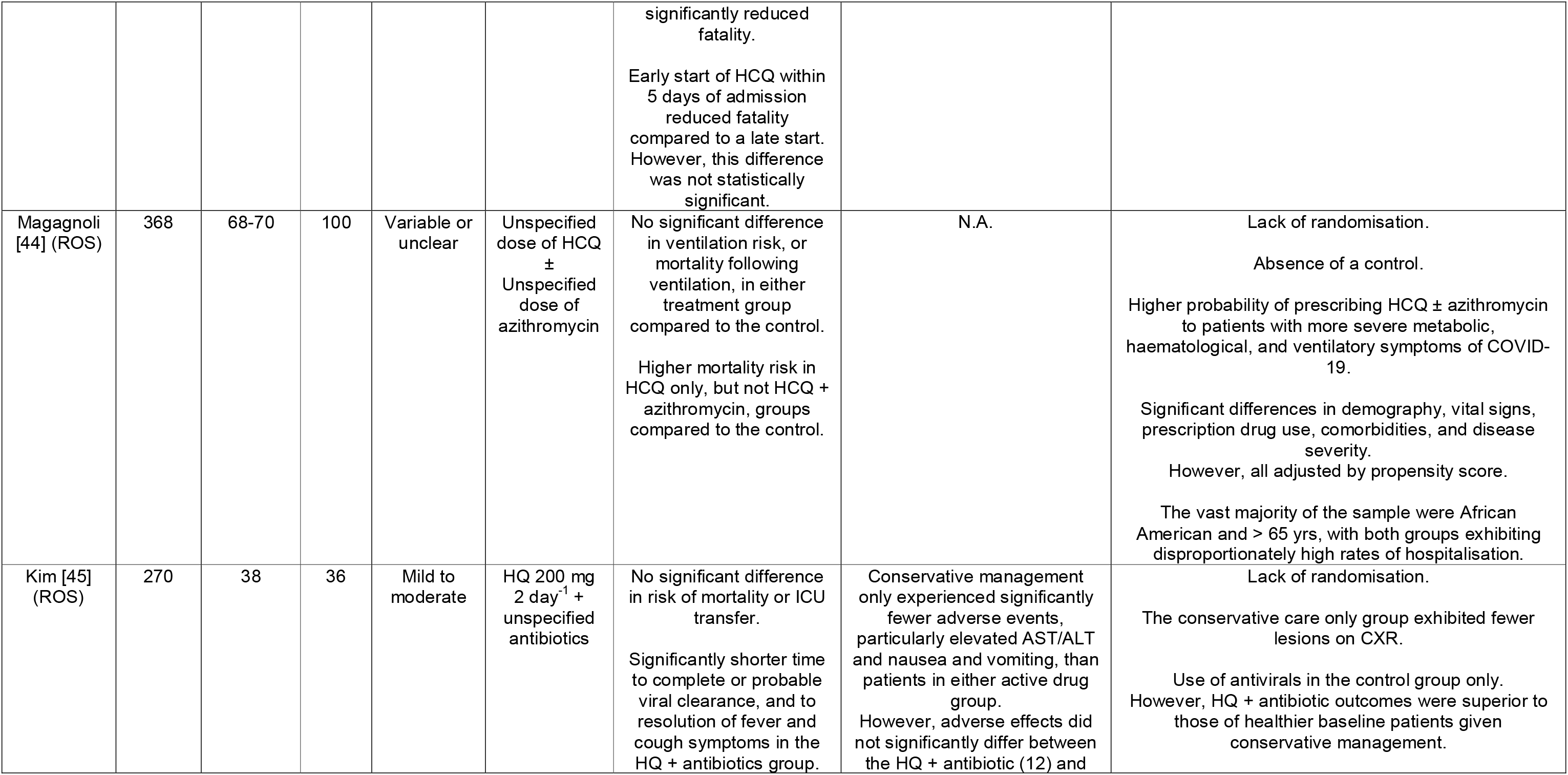
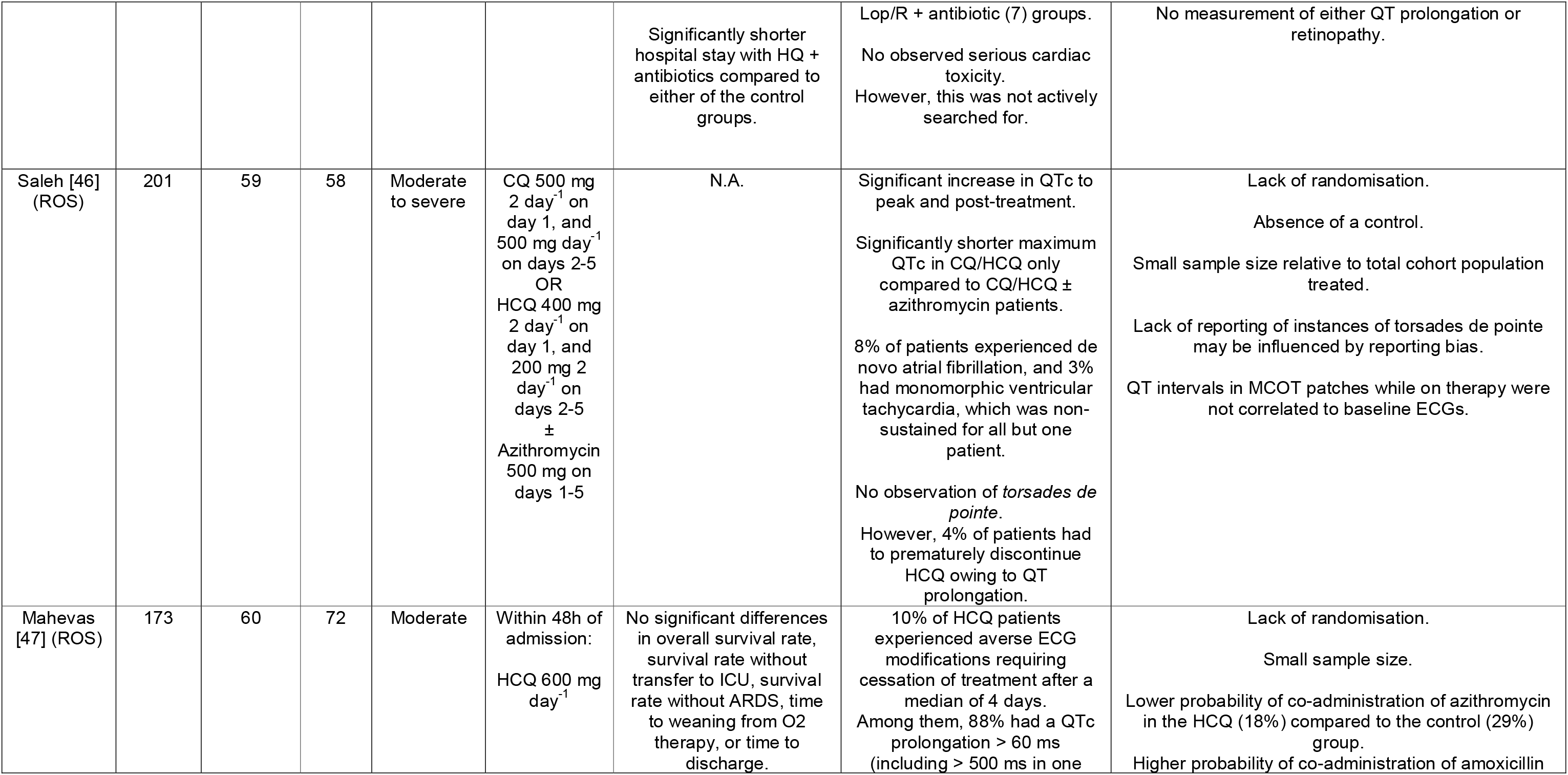
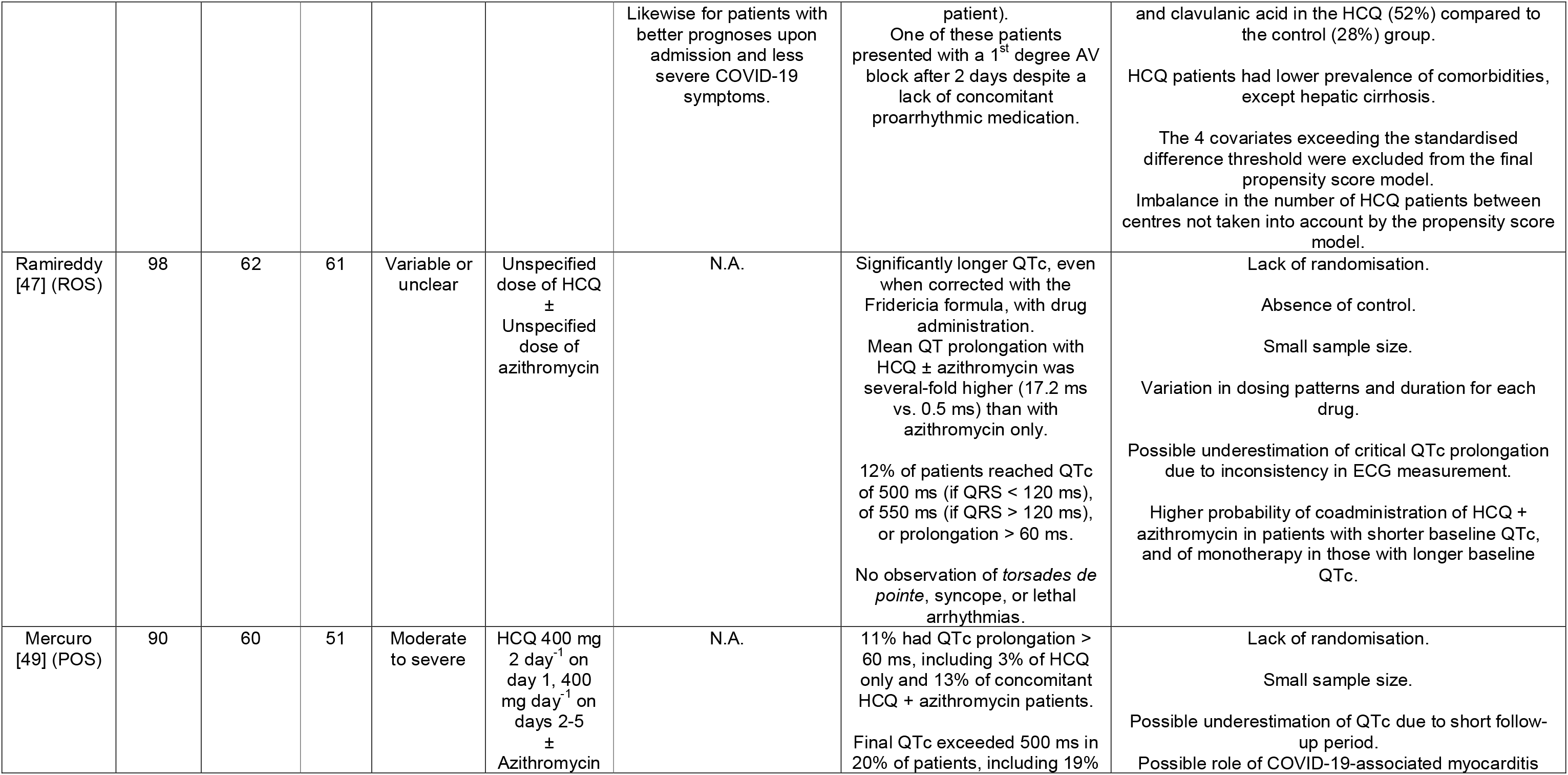
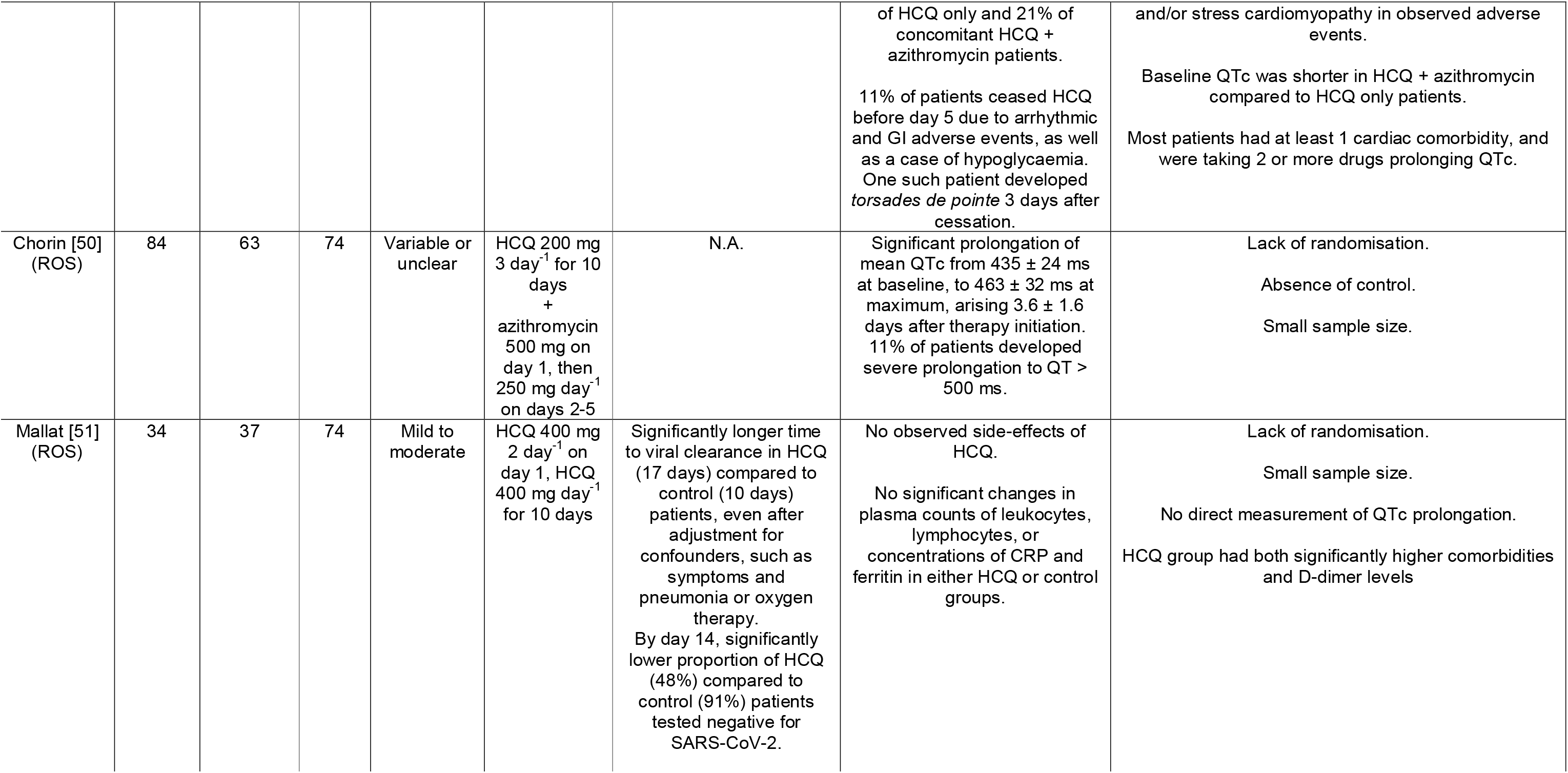
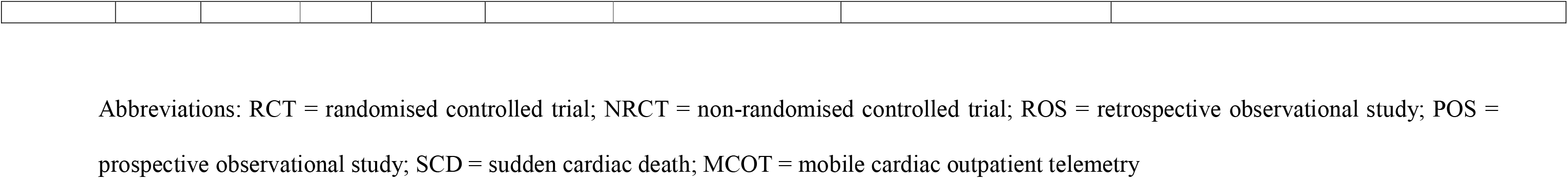
Data extraction from the 19 papers passing quality appraisal.

## 3.0 Results

### 3.1 Search Breakdown

The results of each search, and the number failing and passing exclusion and inclusion criteria, respectively, have been summarised in a flowchart (Figure 1).

**Fig. 1:**
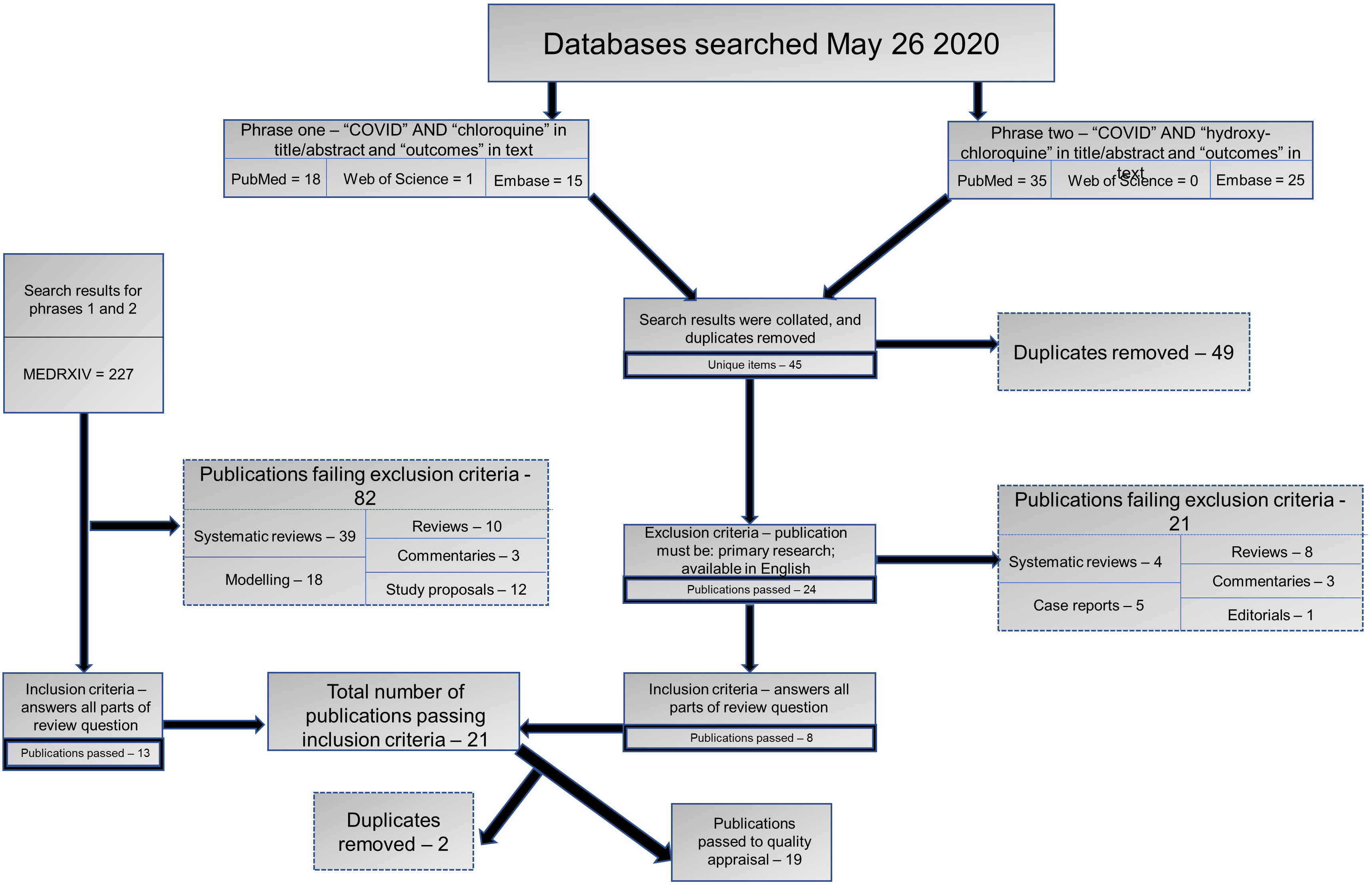
The attrition processes for publication. This comprised: (1) searching of three databases for peer-reviewed papers; (2) acquisition of submitted, but not yet peer-reviewed, items from a preprint server; and (3) application of exclusion and inclusion criteria. The removal of duplicates at each step, where necessary, left 19 unique items to pass onto the quality appraisal stage

On May 26 2020, searches for phrases one (“COVID” AND “chloroquine”[title/abstract] AND “outcomes”[full text]) and two (“COVID” AND “hydroxychloroquine”[title/abstract] AND “outcomes”[full text]) yielded results distributed as follows: 53 on PubMed; one on Web of Science; and 40 on Embase. The subsequent removal of 49 duplicates left 45 unique items from these three databases. In parallel, use of the same stenography in searching MEDRXIV yielded 227 unique results.

Of the unique papers discovered in the first three databases, 21 failed the exclusion criteria, distributed categorically as follows: eight reviews; five case reports; four systematic reviews; three commentaries; and one editorial. Likewise, of those unique items found on the preprint servers, 82 were excluded by the same criteria, distributed categorically as follows: 39 systematic reviews; 18 models of COVID-19 spread and/or symptoms; 12 study proposals and/or protocols; 10 reviews; and three commentaries.

As part of the search attrition methodology, parallel application of the inclusion criteria to each set of remaining unique results left eight and 13 items from the first three, and preprint, databases, respectively. Of these 21 studies, two were duplicates, such that 19 papers answering the research question passed onto quality appraisal.

### 3.2 Quality Appraisal

The use of 13 criteria created in accordance with the aforementioned Checklist of Review Criteria elaborated by the GERA-RIME committee, meeting 12 of which was judged to be indicative of scientific rigour, did not exclude any of the remaining 19 items (Table 2).

### 3.3 Study Design

As a result of the relatively broad scope of the research question, the authors of the 19 papers passing quality appraisal employed a variety of study types, therapeutic doses, and primary and/or secondary outcomes (Table 2). It is thus essential to distinguish results by study design in order to prevent invalid inferences drawn from comparison of data sets.

#### 3.3.1 Study type

Although the research question requires the use of CQ and/or HCQ in patients suspected of, and diagnosed with, COVID-19, there exists a range of possible approaches to the collection of data obtained from such patients.

The gold standard of primary clinical research into the efficacy and safety of drugs administered to humans is the randomised controlled trial, evidence from which may be further buttressed by masking of subjects, experimenters, or both, as well as the use of a placebo in the control group. However, only 21% of items passing quality appraisal were clinical trials, of which, though ¾ were randomised, none employed either masking or an exclusively placebo control. While unacceptable under normal circumstances, the absence of both masking and a placebo is admissible in light of the ethical violation that would otherwise result from the use of either in the context of patient consent being unlikely.

The remaining 79% of papers were observational studies, of which 93% retrospectively searched hospital databases to, and 7% prospectively, collect clinical data obtained by following up on cohorts of patients from the time they received CQ and/or HCQ, or the standard of care only, until a defined end-point. Despite the vast majority (73%) using a case-control structure, a minority (27%) constituted case series focusing on the cardiac safety of drugs administered to hospitalised COVID-19 patients for a given duration of time.

#### 3.3.2 Therapeutic doses

The deliberate absence of a specified dose in the research question accounts for the diversity of administration regimens among the 19 papers. Indeed, the doses used largely reflect the studies being conducted on different dates, which, in turn, influences the relative sway of either federal healthcare guidelines or the results of prior clinical research on regimen selection.

Every clinical trial tested a distinct dosing scheme: 500 mg CQ, twice a day for 10 days [36]; 600 mg CQ, twice a day for 10 days [34]; a loading dose of 1200 mg HCQ per day for 3 days, followed by a maintenance dose of 800 mg per day for 2 or 3 weeks if symptoms are mild/moderate or severe, respectively [33]; and 200 mg CQ three times a day for 10 days, with or without 500 mg azithromycin for 1 day, followed by 250 mg per day for 4 days [35]. Likewise, each clinical trial treated its control group differently, from: 400/100 mg lopinavir/ritonavir twice a day for 10 days [36]; to 450 mg CQ and one placebo tablet twice a day for 1 day, followed by 450 mg CQ and one placebo tablet first and 4 placebo tablets second for 4 days, followed by 4 placebo tablets twice a day for 5 days [34]; and standard of care only [33][35].

By contrast, the retrospective and often multi-centre nature of many observational studies has resulted in 40% using variable or undeclared doses, all collecting data from some patients taking azithromycin in combination with the HCQ. The remaining 60% of items relied on highly divergent dosing regimens (Table 2). One popular iteration administered 200 mg HCQ twice a day, along with: undeclared antibiotics [45]; 500 mg azithromycin per day for 1 day, followed by 250 mg per day for 4 days [50][42]; no antibiotics, and no time declaration [43]. The most common higher dose of choice involved giving 400 mg HCQ, twice a day for 1 day, followed either by: 500 mg per day for 10 days [51]; 400 mg per day for 4 days, with or without an undeclared dose of azithromycin [49]; or 200 mg twice per day for 4 days, with or without 500 mg azithromycin per day for 5 days [46]. In contrast, two studies relied on a much higher loading dose of 600 mg HCQ, either once per day [47] throughout, or twice a day for 1 day, followed by 400 mg per day for 4 days, and with or without 500 mg azithromycin per day for 1 day, followed by 250 mg per day for 4 days [41]. Only one set of authors also analysed data for patients taking CQ, at a dose of 500 mg twice a day for 1 day, followed by 500 mg per day for 4 days, with or without 500 mg azithromycin per day for 5 days [46]. Similarly, the 73% of studies with a control treated the group differently, giving standard of care without (55%), or with declaration of additional antivirals (18%), antibiotics (18%), or both (9%).

#### 3.3.3 Primary and/or secondary outcomes

Given the vast array of possible measures of CQ and HCQ efficacy and safety in COVID-19 patients, the different authors outlined distinct primary and/or secondary outcomes.

The primary outcome in 75% of clinical trials was viral clearance, the definition for which varied from C_t_ > 40 (67%) to C_t_ > 35 (33%) for PCR amplification of SARS-CoV-2 RNA. Likewise, 53% of observational studies directly measured a specific indicator of efficacy other than mortality rate, using similar outcomes to the clinical trials, as well as the duration of hospital stay, need for mechanical ventilation, and probability of transfer to an intensive care unit (ICU).

As regards direct measurement of safety, 75% of clinical trials and 73% of observational studies recorded specific adverse events as an indicator of CQ and/or HCQ safety in COVID-19 patients, with 63% of the total actively focusing on cardiac pathology.

Notably, 53% of studies used mortality rate as an end-point. In isolation, however, risk of death could be indicative of either safety or efficacy. As such, this review reports the findings on mortality rate separately from those pertaining to outcomes that are specific measures of one of efficacy or safety.

### 3.4 Results

Of the clinical trials providing data on a specific indicator of CQ and/or HCQ efficacy in patients suspected of, and diagnosed with, COVID-19, 67% showed a significant increase in the probability of viral clearance in the treatment, compared to the control, group [36][35]. Conversely, 33% failed to find any significant difference in the likelihood of negative SARSCoV-2 conversion [33], despite using a larger dose of HCQ in patients, of whom 99% exhibited only mild to moderate symptoms.

In contrast, 75% of observational studies employing an endpoint specific to efficacy recorded no significant difference in the attainment of outcomes, such as duration of hospital stay, need for mechanical ventilation, and probability of transfer to an intensive care unit (ICU), between COVID-19 patients given a range of CQ and/or HCQ doses, and the control groups. One such study, however, discovered a significantly lower mean HCQ dose in patients with better clinical outcomes [42], while the two remaining sets of authors found either a significant deceleration [51] or acceleration [45] in viral clearance, the latter conflicting with its own data on an unchanged probability of ICU transfer.

All clinical trials and 82% of observational studies examining an indicator unique to drug safety discovered a higher probability of adverse events in those treated patients suspected of, and diagnosed with, COVID-19. Seventy-five percent of the total papers focusing on cardiac side-effects found a greater incidence among patients administered a wide range of CQ and/or HCQ doses, with QTc prolongation the most common finding, in addition to its consequences of VT and cardiac arrest.

Of the total studies using mortality rate as an end-point, 60% reported no significant change in the risk of death, while 30% showed an elevation, and 10% a depression, in treated relative to control patients.

## 4.0 Discussion

The absence of a pharmacological treatment tailored to COVID-19 has rendered urgent the search to find alternative therapies by repositioning drugs with the theoretical potential to alleviate symptoms. However, that a solution is hypothetically plausible is insufficient grounds for translation into clinical practice. Indeed, any therapeutic repurposing must only proceed in light of strong evidence for the pre-clinical basis, and clinical efficacy and safety, of the drug in question. This review finds that, while such evidence certainly exists for the former, it does not for the latter, calling into question any clinical use of CQ and/or HCQ in COVID-19 patients in the absence of high-quality randomised clinical trial data.

### 4.1 Pre-clinical indications of the potential of CQ and HCQ to treat COVID-19

Pre-clinical studies performed *in vitro* provide strong evidence for the theoretical utility of CQ and HCQ in inhibiting all stages of viral entry, maturation, and spread.

*In vitro*, CQ blocks infection both at, and after, entry of SARS-CoV-2 into Vero E6 cells, with an EC_50_ of 1.13 μM [19]. Indeed, although therapeutic doses of CQ do not seem to alter S protein glycosylation [52], whose pattern is distinct from that of SARS-CoV-1 [53], they may inhibit biosynthesis of sialic acid [54], *N*-glycosylation of ACE2, as well as downregulating the expression of PICALM [55] in the clathrin-dependent endocytosis machinery.

Furthermore, immunofluorescence analysis of the amount of NP in distinct vesicular compartments of the host cell has demonstrated that treatment of infected cells with CQ and HCQ stalls transfer of viruses from early to late endosomes [56]. In fact, by increasing the pH of the early endosome, CQ has the potential to reduce acid-dependent proteolytic cleavage of the S protein by cathepsin and TMPRSS2, thereby inhibiting viral uncoating, genomic replication and particle maturation [57]. Despite its similar effect on viral distribution, as well as its comparable cytotoxicity [58], to CQ, HCQ appeared to amplify and enlarge the late endosomes, implying a slightly distinct mechanism of action. Furthermore, there exists conflicting evidence for the relative *in vitro* efficacy of the two drugs [59].

It is nonetheless clear that the initial basis for investigating the translatability of CQ and/or HCQ to the treatment of hospitalised COVID-19 patients was predicated on high-quality evidence for its pre-clinical antiviral efficacy.

### 4.2 Clinical evidence of the efficacy of CQ and HCQ in the treatment of COVID-19

That CQ and HCQ can reduce viral entry, trafficking, and budding *in vitro* constitutes evidence of translational potential relies on the underlying assumption that symptom severity is a function of viral replication. Yet, while viral load may influence severity in the very early stages of COVID-19 [60] – as in SARS [61] –, subsequent symptoms result firstly from the initial cytokine wave of the innate immune response [62], then a state of immunodeficiency and lymphopenia [63], and, finally, a potentially lethal cytokine storm [64]. The causal distinction between these symptomatic phases highlights not only the difficulty in repurposing a single drug for use at all time-points, but also the need to approach with caution the comparison of trial data collected from patients given drugs at different times post-infection.

The sample of one of the first clinical trials performed on patients testing positive upon PCR amplification of SARS-CoV-2 RNA comprised asymptomatic patients (17%), as well as those with upper (61%) and lower (22%) respiratory tract infections, thereby capturing the range of symptom severity. After 6 days of treatment, patients given HCQ alone had a higher probability of viral clearance compared to those given the standard of care only (57% vs. 13%), rising to 100% in patients also given azithromycin [35]. That the authors additionally discovered a greater drug effect on patients with upper and lower respiratory tract infections than on asymptomatic individuals raises the possibility that the potential therapeutic benefit of HCQ in COVID-19 patients lies in its capacity for immunomodulation. On a theoretical level, the anti-inflammatory effects of HCQ render such an effect possible. Indeed, through alkalinisation of early endosomes, CQ and HCQ could impair: PAMP-induced activation of TLR7 and TLR9 [65], and, by extension, MMP-9 expression [66]; antigen presentation by major histocompatibility complexes (MHCs) [67][68]; prostaglandin and thromboxane production [69]; and T and B cell activation [70], differentiation, and proliferation [20]. Importantly, both SARS-CoV-2 [71] [72] and related coronaviruses, such as SARS-CoV-1 [73] and MERS-CoV [74], may incur pulmonary damage through TNF-(alpha) [75], which CQ and HCQ can down-regulate through p38 MAPK inhibition [76].

However, this study had several considerable limitations. In addition to the lack of randomisation and the use of a C_t_ of 35 rather than 40 as the threshold for viral clearance, the sample size of 36 was very small. Moreover, 23% of patients in the treatment, but none of those in the control, group were lost in the follow-up due to transfer to the ICU, disenrollment, or premature cessation, leaving the sample even further underpowered. Indeed, Bayesian reanalysis of the data demonstrates that the statistical evidence for efficacy weakens to anecdotally positive upon the exclusion of untested patients, and even to anecdotally negative with the assumption that untested patients were infected with SARSCoV-2 [77].

Other research coming to the same conclusion regarding the efficacy of CQ and/or HCQ in COVID-19 patients similarly exhibited numerous shortcomings. For instance, although a randomised controlled trial found that, compared to the control group administered lopinavir/ritonavir, patients with moderate and severe symptoms given CQ exhibited more than double the rate of improvement in CT scan indicators of pulmonary health, the sample consisted of only 22 individuals [36]. Likewise, half of the observational studies showing a lower mortality rate or higher probability of viral clearance with low dose CQ and/or HCQ treatment in patients with severe symptoms were severely underpowered, with a significant imbalance in size of the treatment (48) and control (502) groups [43].

By contrast, the majority of observational studies failing to find a significant difference in the mortality rate between COVID-19 patients treated with CQ and/or HCQ and those given the standard of care (with or without additional antibiotics or antivirals) were sufficiently powered. Furthermore, the homogeneity of, and correction for, baseline characteristics in the case and control cohorts further buttresses the reliability of the evidence presented by studies with sample sizes of 1,061 [42], 1,376 [41], 1,438 [40], 3,372 [38], and 96, 032 [37]. Although one study with a sample size of 2,512 used a treatment cohort with a significantly lower age but greater symptomatic disease compared to the control [39], the vast majority of papers coming to this conclusion despite a significant difference in sample structure and comorbidities were underpowered and thus unlikely to influence the conclusion of this review [47]. Likewise, the only retrospective cohort study suggesting that HCQ delayed viral clearance in COVID-19 patients with mild to moderate symptoms suffered from both a very small sample size of 34, and considerably more frequent comorbidities in the treatment group.

As such, it is reasonable to conclude that the weight of present evidence does not come down in favour of either CQ or HCQ being efficacious in the treatment of COVID-19 patients, relative to the standard in-hospital management of symptoms. Indeed, even if the evidence demonstrating a lack of efficacy were absent, the poor quality of data suggesting any significant benefit should preclude any definitive judgment until the completion of high-quality randomised controlled trials.

### 4.3 Clinical evidence of the safety of CQ and HCQ in the treatment of COVID-19

Robust evidence for the safety of an otherwise efficacious drug is a prerequisite for its widespread application in any clinical setting. In light of the inefficacy – or, at least, wholly unsubstantiated benefit – of CQ and/or HCQ administration in patients with COVID-19, there exists an even more compelling imperative to ensure that any compassionate use did, and does, not contribute to excess mortality.

As an aminoquinolone, CQ, and its derivative, HCQ, are proarrhythmic [78]. Arrythmias arise from an imbalance of the normal physiological variables influencing the activation and inactivation kinetics of the cardiac ion channels that permit the transmembrane currents forming the foundation of the cardiac action potential. In a healthy *milieu intérieur*, the waveform of this cardiac action potential is quadriphasic [79]. Electrical diastole (IV) precedes a rapid depolarisation (0), after which a refractory period is followed by a slow hyperpolarisation (I), a 300 ms plateau (II), and then a period of repolarisation (III) to resting membrane potential (E_m_). By blocking – in order of increasing potency – the delayed (I_Kr_) and inwardly-rectifying (I_K1_) K^+^ currents [80], and the latter preferentially at depolarised E_m_, CQ and HCQ significantly prolong the QT interval and slow ventricular conduction, thereby predisposing to early-after-depolarisation and, by extension, *torsades de pointe*. Combined with their tonic block of voltage-gated Na^+^ and L-type Ca^2+^ currents at low channel opening frequencies, QT interval prolongation thus significantly increases the risk of potentially fatal VTs.

Indeed, there is considerable evidence that CQ and/or HCQ treatment predisposes COVID-19 patients to tachyarrhythmia. In fact, a significant association of high doses of CQ with lethality in patients with severe symptoms forced the premature termination of a randomised controlled trial [34]. Despite this relationship with mortality risk disappearing upon correction for age, there remained a significantly higher proportion of patients in the high (19%) compared to the low (11%) dose group with QTcF > 500 ms, including 2.7% patients who experienced VT before death. However, given the abortion of the study, as well as the co-administration of QT-prolonging oseltamivir [81] confounding the causal link to cardiac side-effects, these data, alone, are insufficient to conclude that CQ is unsafe in COVID-19 patients.

However, several highly powered retrospective observational studies have found significant excess mortality in patients treated with CQ and/or HCQ relative to controls, providing further evidence of a lack of drug safety. For instance, in 96, 032 patients with no significant differences in comorbidities between groups, patients given CQ (4%) or HCQ (6%) alone had a significantly augmented risk of *de novo* in-hospital ventricular arrhythmias, compared to controls (0.3%) given standard therapy, including remdesivir [37]. Notably, the addition of a macrolide antibiotic further increased VT risk, owing to synergistic prolongation of QTc. Furthermore, ¾ of the remaining studies with smaller cohorts – but still over 1,000 – demonstrated either a significant increase in the risk of VT or of cardiac-related mortality incurred by the administration of CQ. Moreover, despite being individually underpowered, several smaller retrospective database searches focusing specifically on the QTc duration have consistently and independently supported a significant prolongation in hospitalised COVID-19 patients treated with a range of CQ and/or HCQ doses [46][49][50][48].

From a theoretical standpoint, however, the cardiac safety risk of CQ and HCQ use is unlikely uniform among COVID-19 patients. Indeed, there is a significant positive correlation between baseline QTc and age [82], QTc prolongation and antidysrhythmic, antipsychotic or macrolide antibiotic coadministration [83], and QT interval dispersion and mortality risk in type II diabetes mellitus [84]. Given the relative risk conferred by both older age [85] and type II diabetes mellitus [86] on symptom severity and consequent probability of hospitalisation of COVID-19 patients, the data for in-hospital CQ/HCQ safety may not be extrapolable to many infected individuals in the population due to selection bias [87]. Regardless, drawing causal inferences from such observational studies is inadvisable given the lack of randomisation and absence of a placebo in the control groups, leaving the data susceptible to confounders. Moreover, the wide variation in average sample patient age, symptom severity and drug dosing regimen (Table 2) further complicates inferences of reliable agreement between the papers. Yet, for the most highly powered of the studies, tipping point analysis suggested that any confounding factor not taken into account would have to exist with a prevalence of 37–50% to render the observed differences insignificant [34]. Furthermore, the strength of the correlative relationship alone between CQ/HCQ use and cardiac side-effects in hospitalised patients is sufficient to warrant extreme caution when administering them to patients in such a clinical setting.

Therefore, as of 26 May 2020, the strongest available evidence suggests that, relative to standard in-hospital management of symptoms, the use of CQ and HCQ to treat hospitalised COVID-19 patients has likely been unsafe. At the very least, the poor quality of data failing to find any significant changes in the risk of VT should preclude definitive judgment on drug safety until the completion of high-quality randomised clinical trials.

## 5.0 Limitations

Crucial to the understanding of the conclusions drawn in this systematic review is an appreciation of its many limitations, which relate to both the search methodology and data analysis.

Insofar as peer-reviewed publications are concerned, this review searched three databases to yield only 45 unique results, leaving the authors to also seek the findings of 227 papers on a preprint server. Despite facilitating the collection of a more representative sample of current research on the subject in question, the absence of documented expert scrutiny ought to prevent their data from influencing clinical decisions. Nevertheless, to compensate for the lack of peer review, rigorous application of the quality appraisal criteria established by the GEA-RIME committee and the Task Force of Academic Medicine ensured that only data from adequately designed studies were taken into account. Importantly, however, that peer-reviewed journal material was no longer a prerequisite for inclusion may have slightly reduced the effects of positive publication bias [88] on the results of this systematic review.

However, the predominant shortcoming of the review is its inability to completely disentangle the large differences in study design when making comparisons between different data sets from the included papers. Indeed, despite stressing the obvious invalidity of cutting across distinct sample sizes, baseline characteristics, drug doses, and individual limitations, a systematic review, by nature, does exactly that. The categorisation of the results and data analysis by randomisation, COVID-19 symptom severity, and HCQ/CQ dosage constitutes an attempt to reduce this problem of comparative inferences as greatly as possible.

## 6.0 Summary and Conclusions

On March 18 2020, the WHO announced the launch of an international phase III-IV randomised controlled trial, with four arms, measuring the efficacy and safety of: (1) remdesivir; (2) lopinavir and ritonavir; (3) lopinavir, ritonavir, and IFN-(beta); and (4) CQ or HCQ [89]. In the meantime, amidst a dearth of high-quality evidence from randomised clinical trials, the U.S. Food and Drug Administration (FDA) issued an emergency use authorisation of CQ and HCQ in COVID-19 patients [90].

Since then, data from the most robust case-control studies have failed to find any significant efficacy, and, indeed, a notable lack of (particularly cardiac) safety for the use of CQ and/or HCQ to treat hospitalised COVID-19 patients. On May 25 2020, the WHO suspended the fourth arm of the Solidarity Trial, citing these safety concerns.

The urge to begin the Solidarity Trial arose from the understandable, and – as aforementioned – necessary desire to rapidly compensate for the prior and present scarcity of randomised data on the efficacy and safety of CQ and/or HCQ in patients infected with SARS-CoV-2. Yet the clinical knowledge of, and subsequent evidence for, the cardiac side-effects of CQ and HCQ, call into question the scientific prudence of the FDA’s initial decision to authorise, and only belatedly caution against, their use in hospitalised patients already at risk owing to comorbidities. In any case, any resumption in administration of either CQ or HCQ in a hospital setting would require strong substantiation of both their safety and efficacy from high-quality randomised controlled data.

## Data Availability

All made available

## Acknowledgements

N.A.

## Conflict of interest statement

None to declare.

## List of abbreviations

CoV: coronavirus
COVID-19: Coronavirus Disease 2019
CQ: chloroquine
FDA: Food and Drug Administration
HCQ: hydroxychloroquine
ICU: intensive care unit
MERS: Middle East Respiratory Syndrome
PICOT: Population, intervention, comparison, outcome, time
PRISMA: Preferred Reporting Items for Systematic Reviews and Meta-Analyses
QTcF: The corrected QT interval by Fredericia
SARS: Severe Acute Respiratory Syndrome
WHO: World Health Organisation
VT: ventricular tachyarrythmia

## Notes

### Competing Interest Statement

The authors have declared no competing interest.

